# Is WHO International Standard for Anti-SARS-CoV-2 Immunoglobulin Clinically Useful?

**DOI:** 10.1101/2021.04.29.21256246

**Authors:** Krzysztof Lukaszuk, Jolanta Kiewisz, Karolina Rozanska, Amira Podolak, Grzegorz Jakiel, Izabela Woclawek-Potocka, Lukasz Rabalski, Aron Lukaszuk

**Affiliations:** Department of Obstetrics and Gynecology Nursing, Medical University of Gdansk, Gdansk, Poland; Invicta Research and Development Center, Sopot, Poland; Department of Human Histology and Embryology, Medical Faculty, University of Warmia and Mazury in Olsztyn, Olsztyn, Poland; 1st Department of Obstetrics and Gynecology, The Center of Postgraduate Medical Education, Warsaw, Poland; Department of Gamete and Embryo Biology, Institute of Animal Reproduction and Food Research, Polish Academy of Sciences, Olsztyn, Poland; Laboratory of Recombinant Vaccines Intercollegiate Faculty of Biotechnology University of Gdansk and Medical University of Gdansk, Gdansk, Poland

**Keywords:** COVID-19, SARS-CoV-2, vaccination, Pfizer/BioNTech, Roche, DiaSorin

## Abstract

**Background:** The introduction of vaccination against SARS-CoV-2 infection needs precise instruments for quality control of vaccination procedure, detection of poor immunological response and estimation of the achieved protection against the disease but also against infection and being infective.

**Objective:** To compare new automated SARS-CoV 2 Ig assay performance characteristics from the automated Elecsys SARS-CoV-2 S (Roche) with the new LIAISON® SARS-CoV-2 TrimericS IgG (DiaSorin) assay and their compatibility with WHO International Standard for anti-SARS-CoV-2 immunoglobulin. In the context of the mass vaccination programs, we undertook the investigation of clinical utility of the two new automated assays by analyzing results in samples collected at specified time points relative to the vaccination time.

**Design:** Prospective assay evaluation.

**Patients:** Medical staff undergoing vaccination with BioNTech/Pfizer Comirnaty vaccine between January and March 2021 (n = 79) and referred for serum antiSARS-CoV 2 Ig testing prior to vaccination, 21 days after the first dose, and 8, 14 and 30 days after the second dose. Main Outcome Measure(s): Serum antibody levels measured with Roche and DiaSorin assays.

**Results:** Intra-assay imprecision was low with DiaSorin at 3.46%; and Roche at 2.5%. The Passing-Bablok regression equation for all tested samples was y (DiaSorin) = 184.61 + (1.03 x Roche) and the correlation between the assays (r=0.587; p < 0.0001).

**Conclusions:** The novel automated assays exhibit strong concordance in calibration, with assay-specific interpretation required for routine clinical use. These results highlight the need for further work on the international standard of measurement of SARS-CoV 2 Ig especially in era of vaccination. The serological assays can be useful to detect IgG/IgM antibodies, to assess the degree of immunization, to trace the contacts, and to support the decision to readmit people to work or vaccinate them again. However, the values generated by both assays can be markedly different, and assay-specific and personalized interpretation is required.

## Introduction

The vaccination against severe acute respiratory syndrome coronavirus 2 (SARS-CoV-2) infection is rapidly changing the epidemic situation in countries undertaking mass vaccination programs. Therefore, it is now important to define and refine vaccination assessment methods to determine the vaccination course and use them to optimize epidemic management protocols. A variety of kits are used for testing the level of antibodies in blood with ELISA methods. Various viral proteins and different classes of antibodies are tested. Concerning the quality of the technology itself and the dependence of its results on a large number of variables, including the influence of the human factor, it is an appropriate technology for initial measurements for the purpose of scientific research. For clinical applications, IVD kits are required - simple to perform (using automatic platforms) and resistant to the influence of human factors. The LIAISON® SARS-CoV-2 TrimericS IgG (DiaSorin, USA) (DiaSorin) assay and Elecsys Anti-SARS-CoV-2 S (IgG and IgM) (Roche Diagnostics, Germany) (Roche) quantitative kits are currently available. The DiaSorin assay detects IgG against the anti-Trimeric Spike glycoprotein of SARS-CoV-2. Roche Ig detection assay is focused on the RBD domain of S1 protein. Both of the assays are potentially useful and easy to use. Recently, both of them were compared by manufacturers to the WHO International Standard for anti-SARS-CoV-2 immunoglobulin [1,2]. The intended use of the International Standard is for the calibration and harmonization of serological assays detecting antibodies neutralizing SARS-CoV-2 [3].

This aim of the study was to assess unification of the anti-SARS-CoV-2 test results by converting them into units of the WHO standard (BAU/ml) and, to evaluate their clinical value for confirmation of prior infection history and detection of suboptimal reaction to vaccination.

The study has been financed by Invicta Research and Development Center.

## Materials and Methods

### Ethical policy

The ethical approval was received from the Ethics Committee at the Gdansk Regional Medical Board (No KB - 4/21). All participants gave written informed consent for providing blood samples.

### Participants

Study included 79 randomly selected participants who underwent SARS-CoV-2 vaccination between Jan. 4, 2021 and Mar. 11, 2021. Participants were divided into two groups based on their status of the SARS-CoV-2 infection prior to vaccination, confirmed by the positive result of antibody test before the first dose of the vaccine. The patients with the history of SARS-CoV-2 (SCV2-positive; n= 15; age: 37.8 (6.13); F/M: 11/4) reported having the infection at least 90 days before vaccination. The SARS-CoV-2 negative patients (SCV2-negative; n= 64; age: 41.54 (11.31); F/M: 55/9) have not reported having contact with a SARS-CoV-2 antigen before vaccination. None of the participants reported any allergic reactions or immune disorders during the vaccination enrollment and medical examination and all received two 30 ug doses of mRNA vaccine (Comirnaty; BioNTech/Pfizer) with the interval of 21 days between doses. Blood were collected from participants before the first dose, after 21 days (i.e., on the day of the second dose) and 8, 14 and 30 days after the second dose. Samples were handled according to the recommendations of the producent of the tests.

### Assays characteristics

Manufacturer-reported assay characteristics, measuring ranges, analytic sensitivity, and detection limits are provided in Table 1.

**Table 1.** The assays characteristics

| Characteristics | Elecsys Anti-SARS-CoV-2 S | LIAISON® SARS-CoV-2 TrimericS IgG |
| --- | --- | --- |
| **Specified by manufacturer:** |  |  |
| Assay type | Automated |  |
| Dilution method | Automated, 10x | Manual, 10x |
| Testing time | 18 min | 35 min |
| Test principle | Double-antigen sandwich principle.<br>Electrochemiluminescence detection (ECLIA) | Indirect immunoassay.<br>Chemiluminescence detection (CLIA) |
| Calibration | 2 points |  |
| Calibrators frequency | Started with new reagent lot / Quality control findings outside the defined limits |  |
| Traceability | Standardized against the internal Roche standard for anti-SARS-CoV-2 S / from date 12.01.2021 Standardized against the WHO IS: NIBSC 20-136 | Correlation with Microneutralization Test (MNT) / from date 02.02.2021 standardized against the WHO IS: NIBSC 20-136 |
| Sample material | Serum |  |
| Sample volume | 12 uL | 10 uL |
| Limit of detection (LoD) | 0.35 U/mL | 0.712 AU/mL |
| Limit of quantification (LoQ) | 0.40 U/mL | 1.63 AU/mL |
| Measuring range | 0.40-250 U/mL | 1.85-800 AU/mL |
| BAU conversion | $\frac{U/mL}{0.972}$ | AU/ml * 2.6 |
| **Laboratory specific:** |  |  |
| Controls frequency | Daily |  |

Two assays were used to determine antibody levels: Elecsys Anti-SARS-CoV-2 S (Roche) and LIAISON® SARS-CoV-2 TrimericS IgG assay (DiaSorin). The Roche assay detects antibodies (IgG) to the SARS-CoV-2 spike (S) protein receptor binding domain (RBD) in human serum and plasma. The DiaSorin assay identifies IgG antibodies against the S1 and S2 proteins of the SARS-CoV-2 virus. Both tests were validated against the WHO International Standard for anti-SARS-CoV-2 immunoglobulin. The quantity of antibody in a tested samples were quantified in the units specific for each assay (U/ml for Roche and AU/ml for DiaSorin) and were converted to Binding Antibody Units (BAU)/ml according to the manufacturers’ information regarding the WHO Standard. The conversion for Roche and DiaSorin tests were: U/ml = 0.972*BAU and AU/ml * 2,6 = BAU/ml, respectively. The results were analyzed for all the collected samples and then separately for the different clinical situations and time points.

When the upper estimating range was acquired, serum samples were diluted automatically (limited to 10x) using the reagents supplied by the manufacturer (Roche), or manually (no need for dilution more than 10x) with antibody-free serum (DiaSorin) and analyzed again. The level of antibodies in the blood of the SCV2-positive group after the first dose of vaccine exceeded the detection range of the Roche kit, and the blood samples from these patients at the post-vaccinations time-points were excluded from subsequent comparisons. Thirty-three samples (51.56 %) from the SCV2-negative group tested 8 days after the second vaccine dose exceeded the detection limit of the Roche kit, thus, the results were excluded from further analysis. The results of the remaining 31 patients of the SCV2-negative group were within the detection range of the Roche kit and were used for comparison.

### Statistical analysis

Statistical analyses were performed using R the packages (tidyverse, mcr) [4,5,6]. Passing-Bablok regression equations were used to estimate the relationship between the results obtained with different analyses. Bland-Altman plots were used to compare tests graphically to assess bias and check whether the variability in measures was homoscedastic. Correlations among the anti SARS-CoV-2 immunoglobulin level in-between conducted tests were evaluated by the Pearson’s rank correlation test.

## Results

### Assay precision and accuracy

The precision and accuracy of assays are shown in Table 2. The obtained imprecision rates (I) for compared sets of analytic and evaluated samples Ig values were satisfactory and did not exceed 4%.

**Table 2.** The precision and accuracy of the ELISA assays

| Control sample | Roche (U/ml) |  | DiaSorin (AU/ml) |  |
| --- | --- | --- | --- | --- |
|  | PreciControl Anti-SARS-CoV-2 S p1 | PreciControl Anti-SARS-CoV-2 S p2 | SARS-CoV-2 TG Control Set p1 | SARS-CoV-2 TG Control Set p2 |
| LOT | 526346 | 526347 | 311031 | 212031 |
| Nominal value | n/a | 8,41 | n/a | 37,5 |
| Range | 0.000-0.399 | 5.887-10.933 | 0.0-6.0 | 26.38-48.8 |
| Average | <0.400 | 7.86 | <1.85 | 38.29 |
| SD | n/a | 0.193 | n/a | 1.323 |
| I (%) | n/a | 2.5 | n/a | 3.46 |
| B (%) | n/a | -6.6 | n/a | 2.1 |

SD – Standard Deviation; I – Imprecision; B - Bias

### Detection of SARS-CoV-2 antibody within the entire study group

Figure 1 presents the Passing-Bablok regressions and Bland-Altman plots comparing Ig values of the same samples, collected before the first dose of the vaccine (n=15), after 21 days (n=64) and 8 (n=31), 14 (n=31) and 30 (n=31) days after the second dose, measured with Roche and DiaSorin ELISA tests. The Passing-Bablok linear regression shows correlation between the assays (r=0.587; p < 0.0001). A disparity was apparent in many results. The Bland-Altman plots show that as the mean value of Ig levels increases, the differences between the two values become larger.

**Figure 1.**
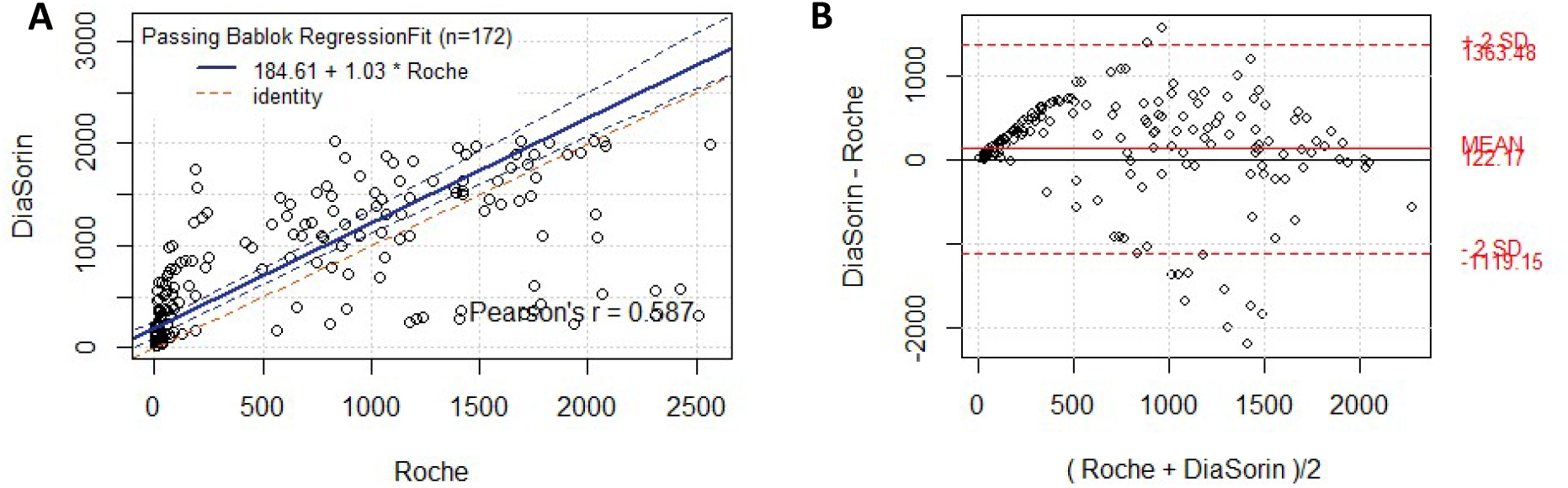
Passing-Bablok and Bland-Altman plots for the same samples compared with Roche and DiaSorin ELISA tests, collected before the first dose of the vaccine (n=15), after 21 days after first dose (n=64) and 8 (n=31), 14 (n=31) and 30 (n=31) days after the second dose, measured with Roche and DiaSorin.

### Detection of SARS-CoV-2 antibody within the SCV2-positive group

The Passing-Bablok regressions and Bland-Altman plots comparing the same samples tested with Roche and DiaSorin ELISA tests for SCV2-positive (Figures 2A and 2B) are presented. For the SCV2-positve group, only samples collected before vaccination are included (n=15), as the results in this group following vaccination were outside the detection limit for the Roche kit. In the SCV2-positive group of patients who suffered from COVID-19 approximately three to six months prior to vaccination, the correlation was found between tested results (r=0.535; p = 0.0399).

**Figure 2.**
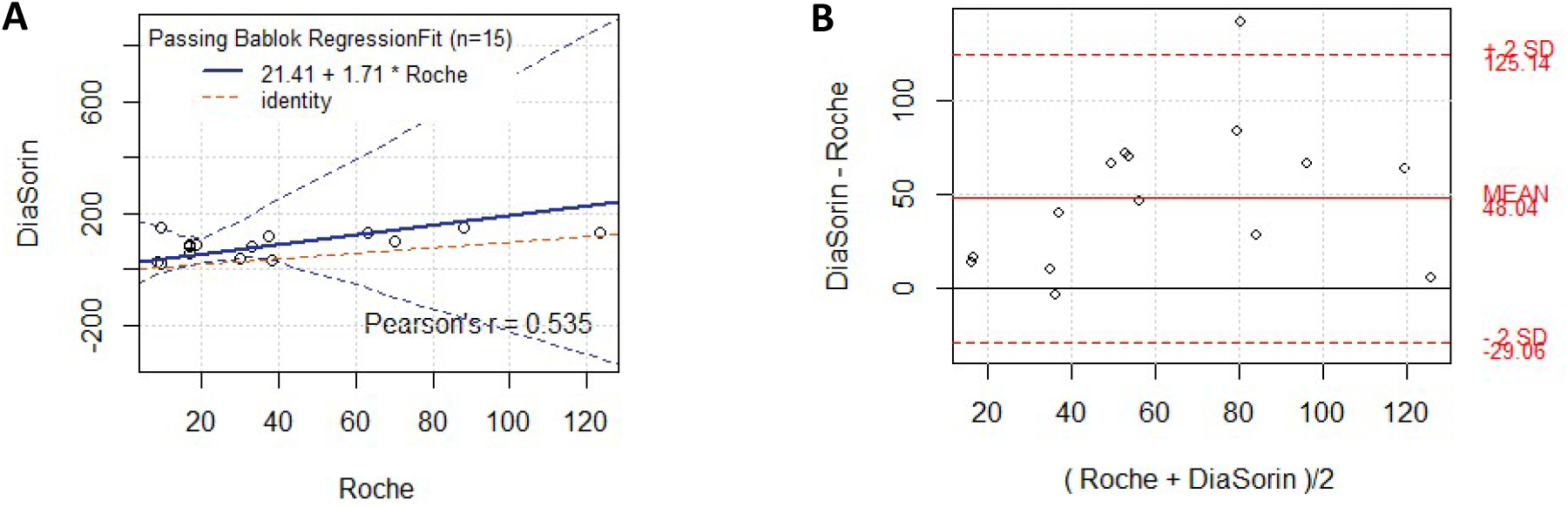
Passing-Bablok and Bland-Altman plots samples from SCV2-positive collected before the first dose of the vaccine (n=15), compared with Roche and DiaSorin ELISA tests.

### Detection of SARS-CoV-2 antibody within the SCV2-negative group prior to second vaccine dose

Passing-Bablok (Fig. 3A) and Bland-Altman (Fig. 3B) plots for the samples from the SCV2-negative group collected on the day of the second vaccine dose (n=64) and measured with Roche and DiaSorin tests are presented. For SARS-CoV-2 antibody level, medium correlation was observed between compared results of the same samples measured with different diagnostic tests (r = 0.425; p = 0.0005).

**Figure 3.**
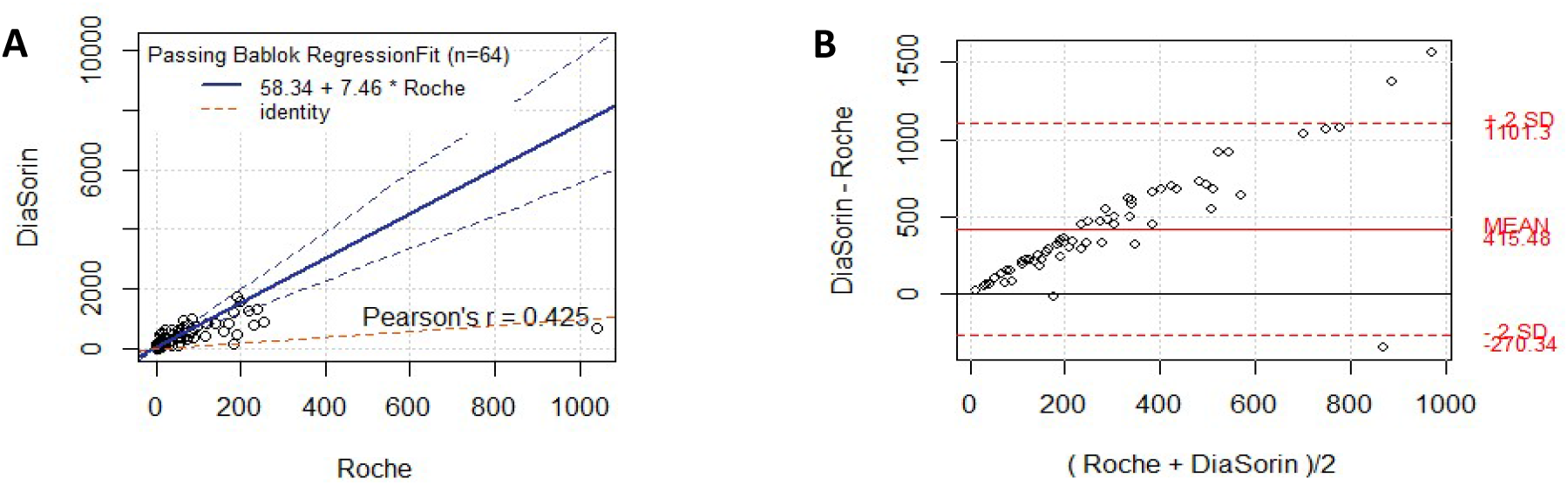
Passing-Bablok and Bland-Altman plots for SARS-CoV-2 antibody level in the SCV2-negative group on the day of the second vaccine dose.

### Detection of SARS-CoV-2 antibody within the SCV2-negative group on day 8, 14 and 30 after the second dose

Following the second vaccine dose, all patients had their SARS-CoV-2 antibody levels measured on day 8, 14 and 30 after the second dose. Only patients whose results remained within the Roche assay detection limit were included in further comparisons of the result obtained on day 8 (n=31), 14 (n=31) and 30 (n=31). Figure 4 shows Passing-Bablok regression (Fig 4 A, C, E) and Bland-Altman plots (Fig 4 B, D, F).

**Figure 4.**
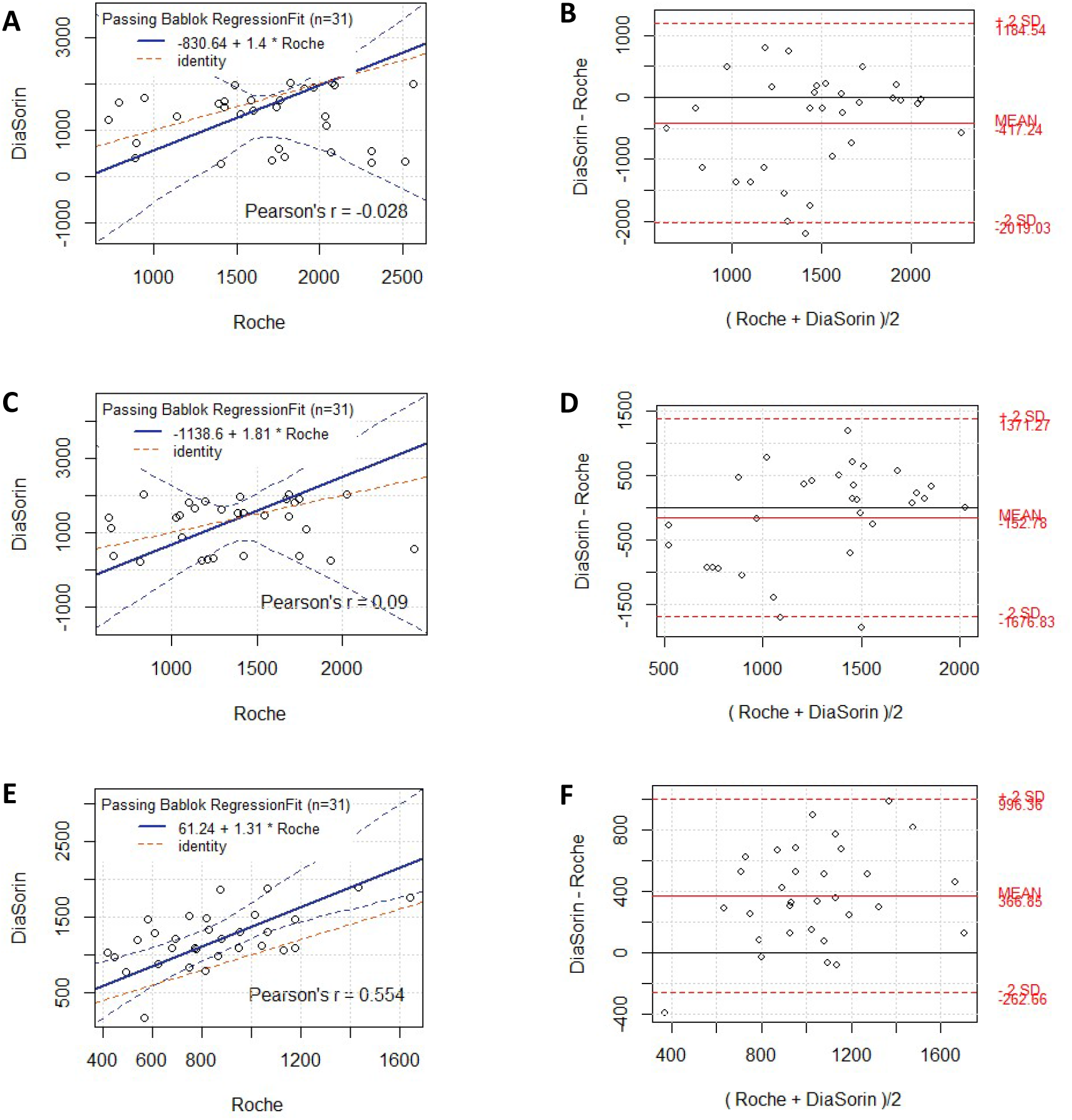
Passing-Bablok and Bland-Altman plots for the subgroup of the SCV2-negative group that remained within the detection range.

Samples tested 8 (Fig. 4 A, B) and 14 (Fig. 4 C, D) days after the second dose of vaccine present very low correlation of test results (r = 0.0284, p = 0.879; r = 0.090, p = 0.629, respectively). The correlation of results of SARS-CoV-2 Ig levels was higher for samples taken 30 days after the second vaccine dose (r = 0.554, p = 0.0012; Fig. 4 E, F).

The dynamics of changes in the antibody levels in tested samples measured with Roche and DiaSorin tests are shown in Figure 5. The mean result for Roche and DiaSorin tests between the samples collected on the day of the second dose and 8 days after the second dose was higher and statistically significant (for both test p < 0.0001). However, differences between the results obtained on day 8 and day 14 (p < 0.0001), and day 14 and day 30 (p < 0.0001), were higher and statistically significant only for the Roche assay. In case of the DiaSorin results the differences were not statistically significant (8/14 day p = 0.7171; 14/30 day p = 0.85).

**Figure 5.**
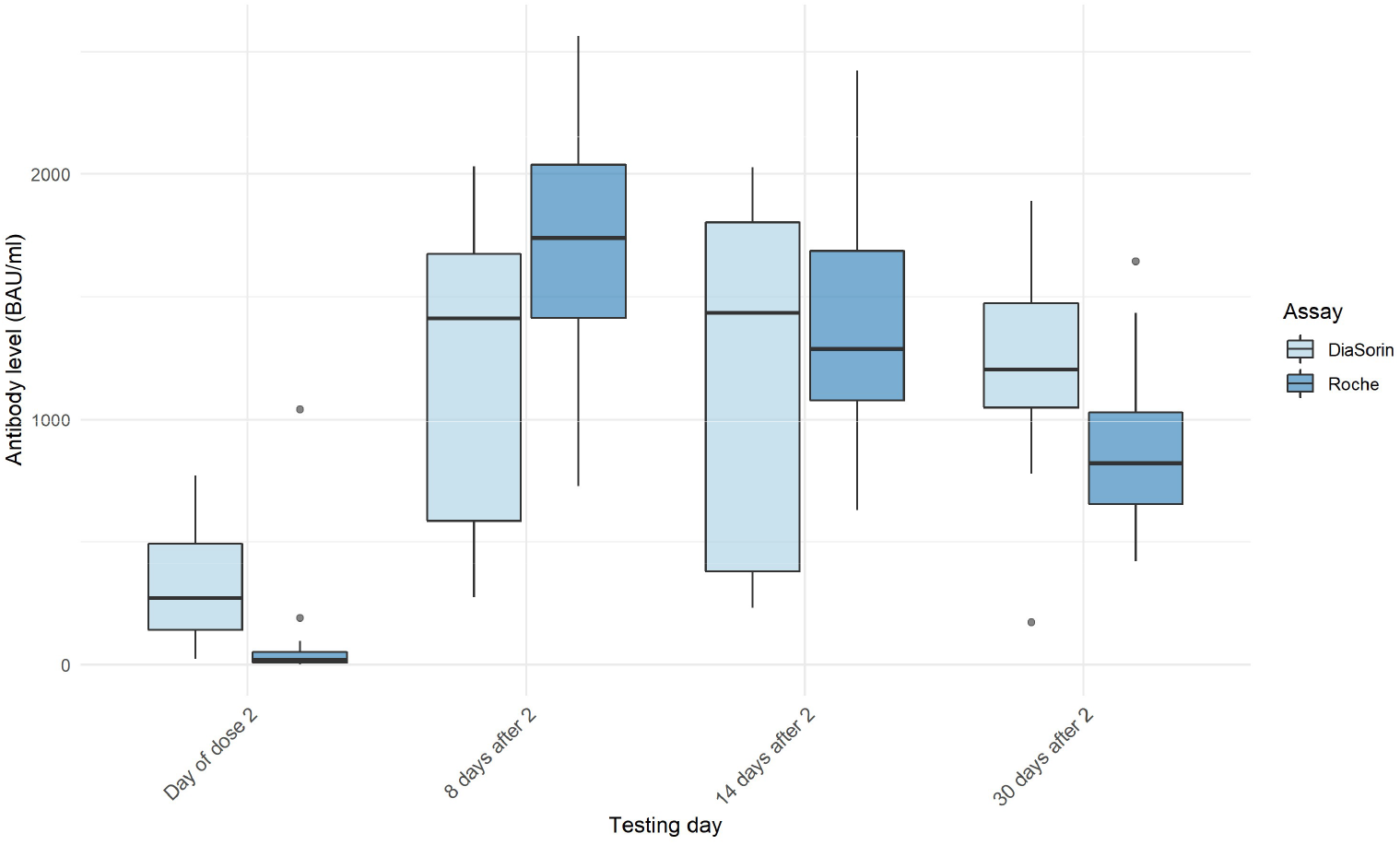
The SARS-CoV-2 antibody levels at SCV2-negative group measured with the Roche and DiaSorin assays at the day of the second dose and 8, 14 and 30 days of after the second dose of the vaccine.

Figure 6 presents the dynamics of changes of the SARS-CoV-2 antibody levels in the SCV2-negative group participants whose results remained within the Roche detection limit (n=31). The profile of the SARS-CoV-2 antibody levels in samples measured with the Roche test shows an expected rising of the antibody level 8 days after the second dose and a steady decline 14 and 30 days after vaccination. Measurement of the SARS-CoV-2 antibody levels with the DiaSorin test shows diversified profile and heterogeneous dynamics of antibodies changes in the tested samples.

**Figure 6.**
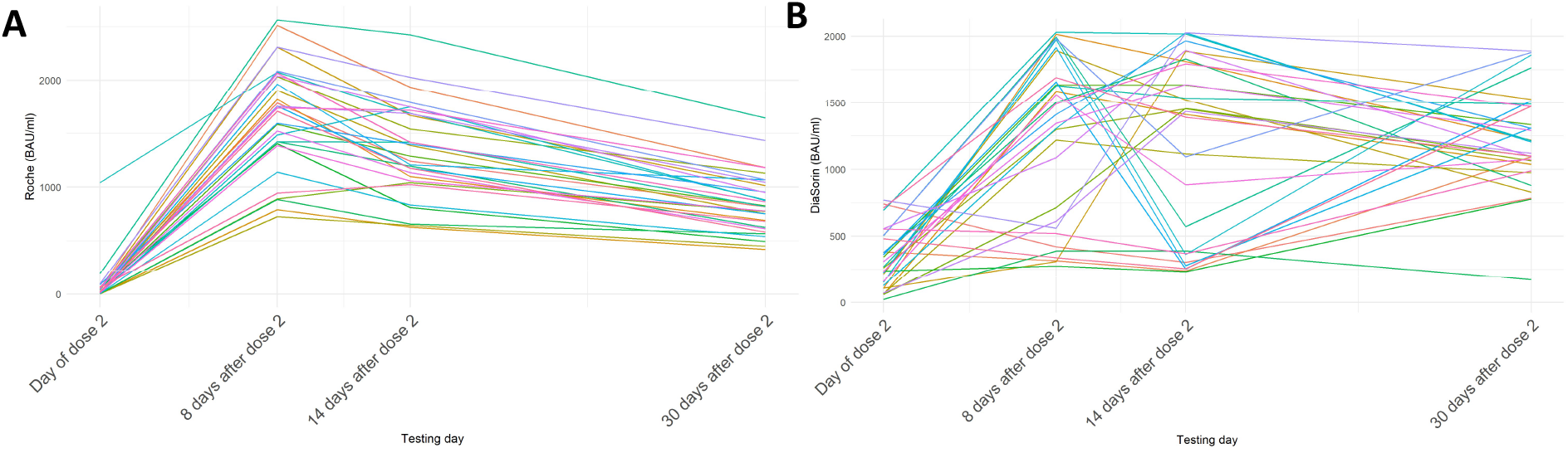
Dynamics of antibody level changes for individual participants as measured by Roche (A) and DiaSorin (B) assays.

## Discussion

The prevalence and high mortality rate associated with SARS-CoV-2 has changed the epidemic, economic, medical, psychological and sociological situation of entire populations. Available vaccines are based on different technologies and will vary concerning the induced immune responses. It is important to validate available methods that can be used to detect previously infected individuals and analyze the response to vaccines. Thus, we examined two IVD kits from major analytical solutions providers (DiaSorin and Roche kits). We confirmed high precision and accuracy of the assays, however, we did observe an increasing difference between the two values when the mean of Ig levels is growing. After division into the SCV2-positive group (confirmed infection approximately three to six months prior to vaccination) and SCV2-negative group, we observed that for the SCV2-positive group the only results that remained within the detection limit for the Roche kit were those obtained before the first dose of the vaccine. In the SCV2-negative group all patients had a result within the Roche detection limit only on the day of the second vaccine dose. Moreover, we found a medium correlation in the SCV2-positive and SCV2-negative group between the Roche and DiaSorin results. When analyzing results that remained within the Roche detection limit, we determined that the correlations dropped down to low for days 8 and 14 and rose to medium for day 30 after the second dose of the vaccine in SCV-negative group. The dynamics of changes of the SARS-CoV-2 antibody levels in SCV2-negative group appears homogenous based on the Roche results but heterogenous when considering the DiaSorin result.

The evaluated Roche and DiaSorin kits are of very high quality, require small sample volumes and have short testing times (12ul and 10ul, and 35 minutes and 18 minutes, respectively). Both kits appear to be very useful in assessing the immune response of vaccinated individuals. The Roche kit makes response assessment difficult due to calibration at a relatively low level and technically unfeasible sample dilution above 10x (dilution over 10x leads to unreliable results - unpublished data). The DiaSorin kit has a very wide measurement range and practically every antibody level produced in a vaccinated person falls within the range that can be determined. What is surprising, however, is the large variability in antibody levels when assessing the dynamics of their changes in individual patients, contradicting our knowledge and the results of the Roche kit.

Antibody response studies to date have mainly relied on results from COVID-19 patients and recovered patients. Studies have mainly addressed qualitative issues - the presence of antibodies and the effectiveness of the kits in detecting infection [7]. The Trabaud study evaluated 8 kits, half of which tested total Ig and the others IgG, four of which assessed binding to the N protein of the virus the others the S protein or its RBD epitome. Their reported sensitivity referred to the percentage of positive results in individuals depending on the time since the onset of disease symptoms. Thus, these data were not based on objective and measurable markers of disease. This study did not look at asymptomatic individuals nor has it included an extended follow-up period after contracting COVID-19. Similarly, the Naaber study [8], although evaluating the clinical use of nine tests, is concerned with investigating the utility of tests to detect or confirm an immune response in the PCR positive patients. A study with a similar objective to ours is a comparison of the quality of automated assays from some of the largest suppliers - Abbott, Roche and DiaSorin by Perkmann et al [9]. This well-conducted study of sensitivity, specificity, and other assay quality parameters based on a very large number of negative samples and 65 SARS-CoV-2 RT-PCR confirmed samples measured in median time of 41 after symptoms or RT-PCR confirmation. They found differences in the evaluated assays that would be diagnostically important and would affect their positive predictive value. They concluded that SARS-CoV-2 antibody tests demand a very high specificity due to the low seroprevalences of the disease. However, this study is also difficult to relate directly to our results. This study involved previous sets of both the DiaSorin (SARS-CoV-2 S1/S2 IgG now SARS-CoV-2 TrimericS IgG test) and Roche (Anti-SARS-CoV-2 (against nucleocapsid) now Anti-SARS-CoV-2 S assay) assays. Both assays have changed their test domains to allow for greater ability to measure quantitative responses and to prepare them for measuring vaccine responses. The test antigen in the DiaSorin kit (recombinant Trimeric Spike glycoprotein) now allows measurement of antibodies against different epitopes of the S protein. The Roche kit is focused on detection of antibodies against the receptor binding domain (RBD) of the spike (S) protein. The Pfizer, Moderna and Astra Zeneca vaccines elicit antibodies to the RBD of the spike protein. Thus, this facilitates a more precise evaluation of the response to these specific vaccines but may make it more difficult to evaluate vaccines with a broader spectrum of induced response.

The results of patients who contracted COVID-19 3-6 months prior to vaccination showed complete qualitative agreement. From the results obtained, it can be concluded that the decision on additional vaccination should be based on the level of antibodies 14-21 days after the first dose, and not on the basis of the baseline antibody level.

We have analyzed the antibody levels at specified time points after the second dose of the vaccine. Our goal was to assess if the weak correlation is caused by the IgM antibodies that emerge after the first contact with the antigen and persist for approximately six weeks [10]. However, our results from samples obtained at time points which should have either no IgM (SVC2-positive prior to vaccination) or minimal IgM (SVC2-negative 30 days after the second vaccine dose – 7 weeks after the first dose) showed only medium correlation between the Roche and DiaSorin results. At the same time we have seen the lowest correlation in the results from days 8 and 14 after the second vaccine dose. These results could possibly be explained by the presence of IgM measured by the Roche assay but not DiaSorin.

This study indicates a further need for unification of the standard and cooperation between assay manufacturers to agree on the cross-utility of their tests. Despite manufacturers’ claims about the value of the tests, they require external validation to detect problems that may be difficult to define within a commercial organization. Tests still need to be validated for virus neutralization to assess the usefulness of the test in evaluating patient protection against infection and in evaluating the clinical status of the patient.

In both cases, the companies confirmed that the results for the WHO reference panel samples are as expected and are easily converted to units of BAU/ml of the WHO international standard (using the conversion factor AU/ml * 2.6 = BAU/ml for the DiaSorin kit and Roche U = 0.972*BAU/ml for the Roche kit). The intended use of the International Standard is for the calibration and harmonization of serological assays detecting anti-SARS-CoV-2 neutralizing antibodies. The results obtained do not show any valuable correlation between the tested kits making it impossible to cross evaluate the results between the kits.

Strengths and limitations of the study require consideration. This study represents a pragmatic comparison of Ig measurement methods using an unselected group of patients referred for clinical Ig measurement before, during and after vaccination. The results still need some independent neutralization antibody measurements tests to be performed for their validation.

## Conclusion

In conclusion, we found a weak correlation between both tested assay. The WHO International Standard has not improved the harmonization of serological assays detecting anti-SARS-CoV-2 neutralizing antibodies. However, we have demonstrated that both assays can be used clinically for the detection of infected subjects, the convalescents’ Ig levels and the confirmation of the vaccination quality and the individual immunological response. The greater precision was exhibited by the Roche kit. The DiaSorin test shows a lot more extensive scope of estimations what reduces the need for sample dilution. On the other hand, Roche results appear better suited to the detection of the results, which we expected as a humoral response to infection. However, the values generated by both assays can be markedly different, and assay-specific and personalized interpretation is required.

## Data Availability

N/A

## Notes

### Competing Interest Statement

The authors have declared no competing interest.

### Author Declarations

The ethical approval was received from the Ethics Committee at the Gdansk Regional Medical Board (No KB - 4/21)

